# Diagnostic performance of host protein signatures as a triage test for active pulmonary TB

**DOI:** 10.1101/2023.01.31.23285229

**Authors:** Lisa Koeppel, Claudia M Denkinger, Romain Wyss, Tobias Broger, Novel N Chegou, Jill M Dunty, Kerry Scott, Tatiana Cáceres, Elloise Dutoit, Cesar Ugarte-Gil, Mark Nicol, Eduardo Gotuzzo, Paul L A M Corstjens, Annemieke Geluk, George B Sigal, Emmanuel Moreau, Audrey Albertini, Anna Mantsoki, Stefano Ongarello, Gerhard Walzl, Marta Fernandez Suarez

**Affiliations:** Division of Infectious disease and Tropical Medicine, University of Heidelberg, Heidelberg, Germany; FIND, Geneva, Switzerland; Meso Scale Diagnostics, LLC., Rockville, Maryland, United States of America; Instituto de Medicina Tropical Alexander von Humboldt, Universidad Peruana Cayetano Heredia, Lima, Perú; Division of Medical Microbiology at the University of Cape Town (UCT), Cape Town, South Africa; Division of Infection and Immunity, School of Biomedical Sciences, University of Western Australia, Perth, Australia; German Center of Infection Research, Focus area Tuberculosis, Heidelberg, Germany; DSI-NRF Centre of Excellence for Biomedical Tuberculosis Research; South African Medical Research Council Centre for Tuberculosis Research; Division of Molecular Biology and Human Genetics, Faculty of Medicine and Health Sciences, Stellenbosch University, Cape Town, South Africa; Department of Cell and Chemical Biology, Leiden University Medical Center, Leiden, Netherlands; Department of Infectious Diseases, Leiden University Medical Center, Leiden, The Netherlands; School of Medicine, Universidad Peruana Cayetano Heredia (UPCH), Lima, Peru

## Abstract

The current four symptom screen recommended by the WHO is widely used as screen to initiate diagnostic testing for active pulmonary tuberculosis (TB), yet the performance is poor especially when TB prevalence is low. In contrast, more sensitive molecular tests are less suitable for the placement at primary care level in low resource settings. In order to meet the WHO End TB targets new diagnostic approaches are urgently needed to find the missing undiagnosed cases. Proteomics-derived blood host biomarkers have been explored because protein detection technologies are suitable for the point-of-care setting and could meet cost targets.

This study aims to find a biomarker signature that fulfills WHO’s target product profile (TPP) for a TB screening. 12 blood-based protein biomarkers from three sample populations (Vietnam, Peru, South Africa) are analyzed individually and in combinations via advanced statistical methods and machine learning algorithms. The combination of I-309, SYWC and kallistatin shows the most promising results for TB prediction throughout the datasets meeting the TPP for a triage test in adults from two countries (Peru and South Africa). The top performing individual markers identified at the global level (I-309 and SYWC) were also among the best performing markers at country level in South Africa and Vietnam.

This analysis clearly shows that a host protein biomarker assay is feasible in adults for certain geographical regions based on one or two biomarkers with a performance that meets minimal WHO TPP criteria.

**Abstract Importance:** Tuberculosis (TB) remains a serious worldwide health problem and diagnosis is hampered by the complexity of tests at primary care level in low resource settings or the low accuracy for screening settings. In order to meet the WHO End TB targets new diagnostic approaches are urgently needed to find the missing undiagnosed cases.

This analysis clearly shows that a host protein biomarker assay is feasible in adults for certain geographical regions. We were able to construct an algorithm through statistical methods and machine learning algorithms whose performance meets the minimum of the WHO target product profile criteria. Thus, further work should be addressed at demonstrating that such as assay can be translated into a practical point-of-care test, and to better understand how to address regional differences in biomarker levels and responses.

## Introduction

New diagnostic approaches are urgently needed to find the ‘missing millions’ of tuberculosis (TB) patients who are never diagnosed and reported and to meet WHO End TB targets (1). Molecular tests like Xpert MTB/RIF assay (hereafter referred to as ‘Xpert’) or Molbio Truenat MTB-RIF Dx are more sensitive than smear microscopy, faster than traditional culture methods and have been rolled out around the world (2). However, placement of the instrument at primary care level in low resource settings where the majority of individuals with TB first access care is restricted by its infrastructure requirements and cost (3, 4).

Currently, the WHO-recommended four symptom screen (W4SS) is widely used as a screen to initiate diagnostic testing for pulmonary TB (PTB). However, the performance of such a screen is poor especially when TB prevalence is low, and better solutions are needed (5). A two-stage diagnostic algorithm in which a highly sensitive and moderately specific test, a so-called ‘triage-or rule-out test’, is applied first at peripheral healthcare settings, could be a cost-effective approach to improving diagnostic yield. Such a test would effectively capture most subjects at higher likelihood of having TB and thus decrease the cost of the expensive confirmatory testing. The target product profile (TPP) suggested by the World Health Organization (WHO) for such a triage test specifies a minimum sensitivity of 90% and specificity of 70% relative to the confirmatory test, for identifying active TB disease and discriminating against the absence of disease (with or without TB infection), or respiratory diseases other than TB (ORD)(6).

Proteomics-derived blood host biomarkers have been explored to develop novel diagnostics for TB (7, 8). They are particularly attractive as protein detection technologies suitable for the point-of-care (POC) are well established, and could meet the cost targets put forward by WHO for a viable diagnostic (6). Current lateral flow technology can cover three biomarkers on one cartridge (9-11). Commercial examples of such multiplex antigen tests are the BD Veritor™ SARS-CoV-2 & Flu A+B combo test by Becton Dickinson and Company (12), Status™ COVID-19/FLU Test by Chembio (13), Sofia 2 Flu + SARS Antigen Fluorescent Immunoassay (FIA) by Quidel (14) and multiplex immunoassays for myocardial infarction such as Quidel Triage Cardiac Panel by Quidel (15). Recently a novel lateral flow based multi-biomarker test was reported for quantitative detection of six biomarkers, indicative for the humoral and cellular response upon infection with the *Mtb*-related *Mycobacterium leprae* (16).

An original large-scale proteomic discovery approach using the SOMAscan (SomaLogic, Boulder, CO) discovered a combination of six biomarkers of high promise and identified additional markers with discriminatory power and large median fold changes (8). Separately, an approach by Walzl et al. captured a partly overlapping, separate group of biomarkers that showed promise to be investigated further (17-20).

In this study, we describe the performance of 12 blood-based host protein biomarkers, selected based on these two prior workstreams, individually and in combination using advanced analysis methods and explore whether the required triage test performance can be met with a protein host-marker signature. We further establish concentration ranges, and nominate key protein marker candidates for translation into such a POC test.

## Methods

### Study design and sample collection

Patients aged 18 years or older presenting with signs and symptoms of TB (cough for at least 2 weeks, fevers, weight loss, and night sweats) and able to provide informed consent were enrolled in South Africa (Division of Medical Microbiology at the University of Cape Town), Peru (Instituto de Medicina Tropical Alexander von Humboldt at Universidad Peruana Cayetano Heredia, Lima) and Vietnam (ham Ngoc Thach Hospital, Ho Chi Minh City). Individuals with signs compatible with only extrapulmonary disease and those having received more than 2 doses of anti-TB therapy prior to enrolment were excluded. Basic demographics information was collected, such as age, weight, and gender, and clinical metadata, such as HIV status and, in some cases, CD4 cell counts and viral load. Data were captured through a dedicated, online, password-protected double data entry system. Chest radiographs were performed and interpreted by local radiologists in a subset of patients (Peru and Vietnam). Participants were asked to provide 2 spot sputum samples, serum, and plasma within 2 days for testing, and HIV testing was offered. The reporting of this study followed the STARD guidelines.

### Biomarkers – Index Test

The biomarker candidates were selected in close discussion with investigators from earlier studies (17-20), considering and balancing individual discriminatory performances, median fold changes and feasibility of detecting the marker or markers in a POC test. From the original SOMAscan discovery work (8), kallistatin, SYWC (an interferon-γ inducible Trp-tRNA-synthetase) and complement component 9 (C9) were selected as they were the first markers when ranked by Kolmogorov-Smirnov (KS) statistic and were part of the original 6-marker signature. Serum amyloid A (SAA) and non-pancreatic Secretory Phospholipase A2 (NPS-PLA2) were included due to their large median fold-change and presence among the top-15 markers. From the work of Walzl et al. (19), ferritin, apolipoprotein A1 (ApoA1), CXCL10 or IP-10, CCL1 or I-309 and CXCL9 or MIG were elected. Finally, two previously described markers with diagnostic and treatment monitoring potential were included as point of comparison, namely lipopolysaccharide binding protein (LBP) and C-reactive protein (CRP) (21). CRP is now endorsed by WHO for screening for TB (22) and was among the top-20 markers in the SOMAscan work which made up an optimal diagnostic bio-signature when combined with I-309 as shown by Walzl et al. In a screen of biomarkers correlating with treatment effect, LBP and CRP were among the top markers with the largest average decreases upon TB treatment (23).

### Reference standard and case definitions

Two sputum samples per study participant were obtained and tested by acid-fast staining, liquid culture using mycobacteria growth indicator tubes (MGIT) with a BACTEC 960 instrument (BD Microbiology Systems, Sparks, MD), solid culture with Löwenstein-Jensen medium, and where available, the Xpert®MTB/RIF (Cepheid, Sunnyvale, CA [Xpert]) test. A culture-positive case was defined as a participant with at least 1 culture testing positive for *Mycobacterium tuberculosis* (MTB) in either of the 2 samples in either solid and liquid culture. A culture-negative case was defined as a participant with all negative cultures for MTB (out of 4) and negative cultures in follow-up. Study participants were considered smear-positive, if they had at least 1 positive smear with acid-fast staining. Participants with unclear microbiological diagnosis were excluded from the analysis, including those with missing culture results, those classified as smear-positive but culture-negative, and those only showing growth of nontuberculous mycobacteria culture.

Patients were categorized based on clinical and microbiological results. Patients with positive MTB cultures were diagnosed as definite tuberculosis and subcategorized into smear-positive and smear-negative groups. Participants who were smear and culture negative but responded to empiric tuberculosis treatment were classified as “clinical tuberculosis” negative (CXR may be abnormal or not). Participants who were smear negative, Xpert and culture negative on all sputum samples and who exhibited symptom resolution in the absence of tuberculosis treatment at the 2–3-month follow-up visit were classified as “non-tuberculosis disease.” See the supplementary material Table S8.

All serum samples were obtained at baseline, given a unique barcode, and frozen on site in 0.5-ml aliquots prior to shipment to a central repository. Frozen serum aliquots were sent to Meso Scale Discovery, LLC (MSD) blinded from the central repository.

### MSD U-PLEX assay testing

Custom immunoassay panels for the pre-defined host biomarkers were developed employing a multiplexed sandwich immunoassay format and electrochemiluminescence (ECL) detection and carried out on commercial instrumentation and multi-well plate consumables from MSD (24). The host biomarker panels were developed and optimized for multiplexing on the MSD^®^ U-PLEX^®^ assay platform, and capture antibody arrays were formed according to manufacturer instructions.

The assay components for each panel included a 96-well plate having an array of capture antibodies in each well (generated by the binding of capture antibodies labeled with U-PLEX linkers), a set of labeled detection antibodies for each analyte in the panel (labeled with the MSD SULFO-TAG™ ECL label), an assay diluent, a detection antibody diluent, a wash buffer, an ECL read buffer (MSD Gold Read Buffer A), a calibration standard containing a blend of the target analytes and a set of controls. Blended-analyte diluent-based controls were created for each panel at two concentration levels (high and low). A matrix-based control was also created by screening and pooling MSD-provided human serum samples. Each plate included the calibration standard, the set of controls and the serum samples analyzed in duplicate wells. Additional details are described in the Supplement C1.

To address the wide range of concentrations covered by the targeted biomarkers, the assays were divided into three panels run with different capture antibody arrays and using different sample dilutions. Panel 1 (IP-10, I-309 and MIG) was run using a 1:2 sample dilution, Panel 2 (NPS-PLA2, Ferritin and SYWC) was run using a 1:50 sample dilution, and Panel 3 (ApoA1, C9, CRP, kallistatin, LBP and SAA) was run using a 1:50,000 sample dilution. Based on the use of CRP as a WHO-recommended screening test, the CRP result from Panel 3 is also presented independently as a comparator.

### Statistical analysis

We evaluated the diagnostic performance of 12 host biomarkers for predicting active TB by means of the efficient use of the machine learning algorithms (25) with the programming language python (Version 3.8) and the *scipy* library. The statistical analysis was first performed on each of the biomarkers individually, then on all possible biomarker combinations. We compared the goodness of fit between the combinations by the value of their negative log-likelihood of the fitted binomial model accounting for country and human immunodeficiency virus (HIV) effect. This estimator is a transformation of the maximum likelihood value yet preserving numerical stability. It allows a ranking of the different combinations, because the smaller the negative log-likelihood value the better is the model fit. For each fixed number of biomarkers, we further analyzed the top three performing biomarker combinations, ranked by their negative loglikelihood value, by using a variety of black box machine learning algorithms. Each methodological concept provides different strengths and limitations allowing for more complex analyses. The algorithmic approaches that were evaluated included logistic regression, random forests representing an ensemble method, support vector machines representing a non-probabilistic binary linear classifier, and Naïve Bayes representing a probabilistic classifier.

The aim of this analysis was to investigate how well the algorithms could predict active TB disease, and to identify promising biomarker combinations for further research. As an exploratory analysis, the full dataset was analyzed using 5-fold cross validation in order to make the best use of the limited data available. For each selected biomarker combination and model, we calculated Receiver Operator Characteristic (*ROC*) curves and compared the results with the TPP goal suggested by the WHO of at least 90% sensitivity and 70% specificity against the reference standard (6).

We further analyzed the algorithms’ performance on subsets by country and HIV status.

### Ethics

Ethical approvals for the study were obtained by the Human Research Ethics Committee of the respective sites including the Vietnam Committee of the Ministry of Health, the City of Cape Town as well as the University of Cape Town Human Research Ethics Committee and the Ethics Committee of the Universidad Peruana Cayetano Heredia. The study was undertaken in accordance with the principles of the Helsinki Declaration. Informed consent was obtained from patients who agreed to participate.

## Results

### Study participants

A total of 479 adults were analyzed in a retrospective nested cohort design with 177 definite tuberculosis cases, and 302 of non-tuberculosis disease (Figure 1). The dataset recorded about the same number of individuals in the respective countries with different prevalence across the countries. In Peru about half of the patients (44%) were TB positive, whereas the prevalence was lower in the other countries (34% in both). In South Africa 53% of the patients were HIV positives regardless of TB status, whereas Peru (0.03%) and Vietnam (10%) showed lower numbers of HIV coinfection.

**Figure 1:**
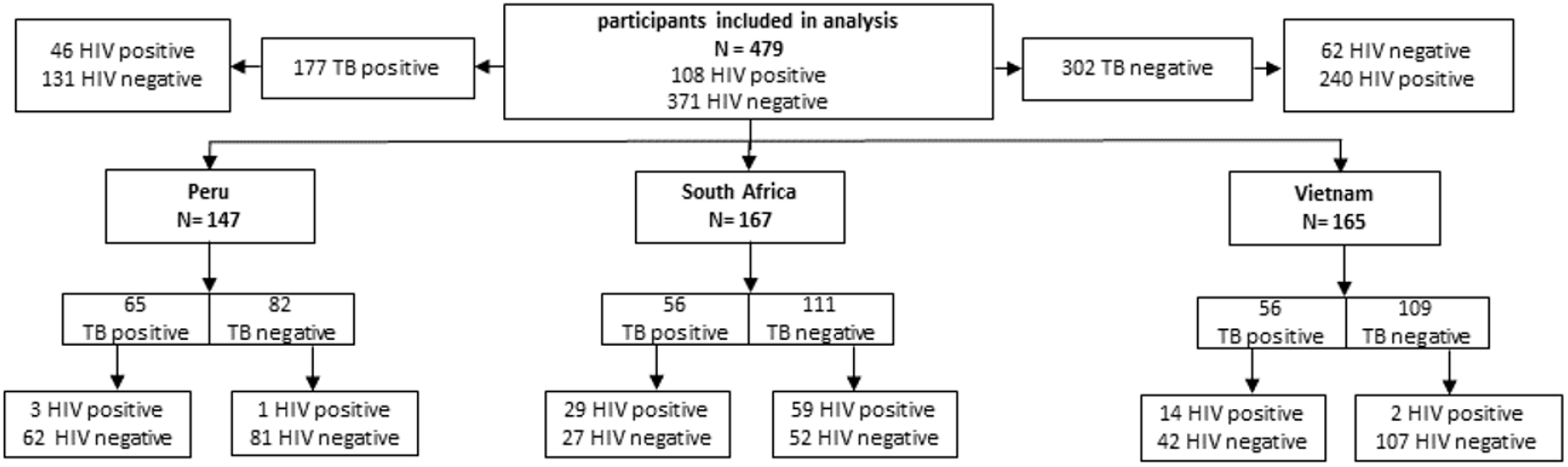
Stratification of the population into country, TB and HIV status

### Host biomarker diagnostic evaluation

Table 1 shows the biomarkers and biomarker combinations ranked by their negative loglikelihood value. For the top performing one, two and three biomarker combinations from Table 1, and for the combination of all 12 biomarkers, Figures 2 and 3 compare the ROC curves generated using four different algorithms, namely logistic regression, random forests, support vector machines and Naïve Bayes. When analyzing single markers, I-309 was top performing according to the value of its negative loglikelihood (area under the curve [AUC] with logistic regression, 0.87), followed by SYWC and MIG (AUC 0.86 and 0.83, respectively). Quantitative information on the host markers’ concentration and fold-changes at global and regional level can be found in Table S4 – S7 in the Appendix Table S4.

**Table 1:** Biomarker combinations ranked according to their value of the negative loglikelihood Interestingly, the biomarker ApoA1 had very poor performance on its own (AUC 0.5; see Appendix), though it seemed to have great additional effect as the second best combination in a subset with three biomarkers ([I-309, SYWC, ApoA1], AUC 0.89).

| **No of biomarkers combined** | **Best combination** | **Second best combination** | **Third best combination** |
| --- | --- | --- | --- |

Table 1: Biomarker combinations ranked according to their value of the negative loglikelihood
|  | [marker(s)]<br>Loglikelihood, AUC<br>for logistic regression | [marker(s)]<br>Loglikelihood, AUC<br>for logistic regression | [marker(s)]<br>Loglikelihood, AUC<br>for logistic regression |
| --- | --- | --- | --- |
| **1** | [I-309]<br>210, 0.87 | [SYWC]<br>213, 0.86 | [MIG]<br>244, 0.83 |
| **2** | [I-309, SYWC]<br>189, 0.88 | [I-309, kallistatin]<br>199, 0.87 | [kallistatin, SYWC]<br>201, 0.88 |
| **3** | [I-309, SYWC,<br>kallistatin]<br>182, 0.90 | [I-309, SYWC,<br>ApoA1]<br>188, 0.89 | [I-309, SYWC,<br>NPSPLA2]<br>189, 0.89 |
| **4** | [I-309, SYWC,<br>kallistatin, C9]<br>179, 0.89 | [I-309, SYWC,<br>kallistatin, NPSPLA2]<br>180, 0.90 | [I-309, SYWC,<br>kallistatin, ApoA1]<br>181, 0.90 |
| **5** | [I-309, SYWC,<br>kallistatin, C9, LBP]<br>174, 0.90 | [I-309, SYWC,<br>kallistatin, SAA, C9]<br>175, 0.89 | [I-309, SYWC,<br>kallistatin, C9,<br>NPSPLA2]<br>175, 0.90 |
| **6** | [I-309, SYCW,<br>kallistatin, C9, LBP,<br>ApoA1]<br>174, 0.90 | [I-309, SYWC,<br>kallistatin, SAA, C9,<br>LBP]<br>174, 0.89 | [I-309, SYWC,<br>kallistatin, C9,<br>NPSPLA2, SAA]<br>174, 0.90 |
| **12** | [NPSPLA2, SYWC,<br>C9, LBP, CRP, SAA,<br>kallistatin, ferritin,<br>IP-10, I-309, MIG,<br>ApoA1]<br>172, 0.89 |  |  |

**Figure 2:**
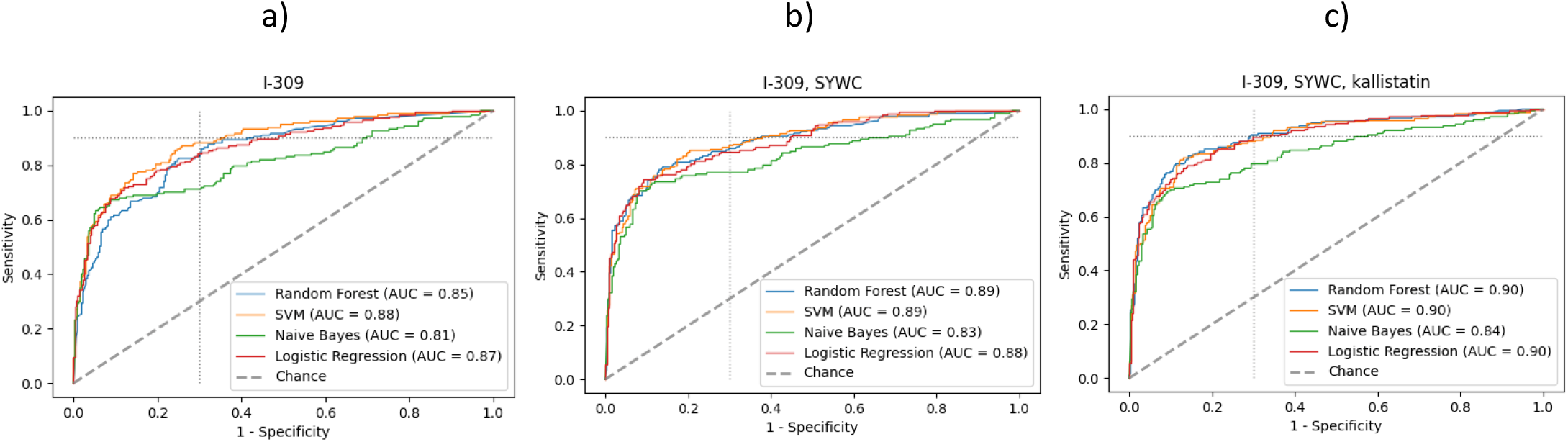
Receiver Operator Characteristic (ROC) curve of different machine learning algorithms with 5-fold cross validation for best performing (a) single biomarker, (b) combination of two and (c) three different biomarkers Caption: The dotted line indicates the minimal TPP target with 90% sensitivity and 70% specificity. With none of the combinations, the TPP could be reached. Abbreviations: SVM: Support Vector Machine; AUC: Area Under the Curve

**Figure 3:**
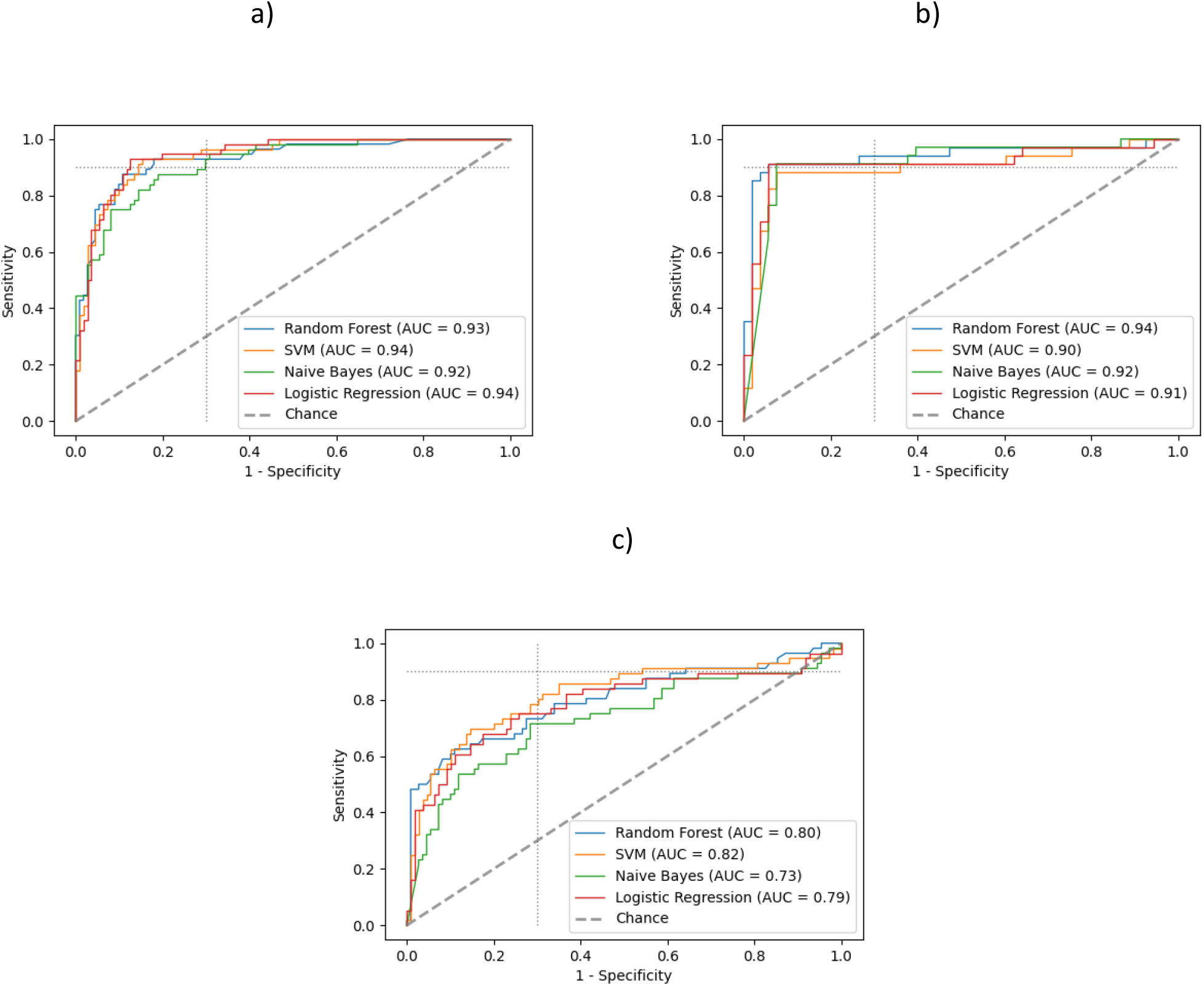
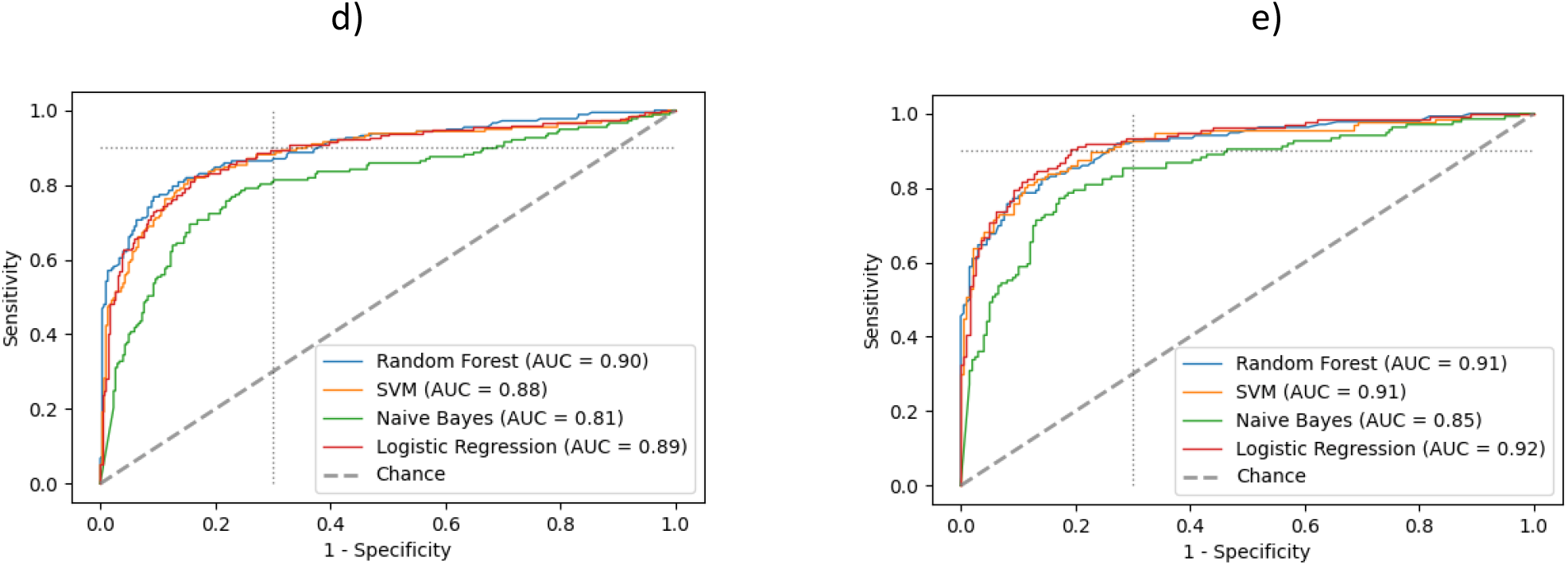
The ROC plot of all 12 biomarkers combined for a) South Africa alone, b) Peru alone, Vietnam alone d) the whole dataset and e) the whole dataset excluding Vietnam. Caption: The dotted line indicates the TPP with 90% sensitivity and 70% specificity. Abbreviations: SVM: Support Vector Machine; AUC: area under the curve

The logistic regression model combining I-309 with SYWC provided a small improvement in the performance relative to I-309 alone (AUC 0.88). The analysis of the ROC curves (Figure 2) for the individual combinations revealed that adding kallistatin as a third marker enhanced the performance in all algorithms confirming the inference (AUC 0.9 for logistic regression compared to 0.87 for I-309 alone). While the Naïve Bayes approach performed poorest (Figure 2c), the triage TPP suggested by the WHO was reached by the random forest algorithm using the combination of I-309, SYWC and kallistatin.

We observe a nested behavior of the top performing biomarker subsets, meaning that the biomarker combinations with less biomarkers were contained in the top performing compounds with more biomarkers. However, combining more than three biomarkers provided no further improvement in the algorithmic performance or the AUC values. For some models we observed a slightly lowered AUC value, in particular when using the full data with 12 biomarkers (AUC decreases for logistic regression to 0.89 in Figure 3d compared to 2c), which can be attributed to overfitting and the randomness of the data sample.

Given the diagnostic performance in the ROC curve and the ranking among the other biomarkers in its group, the combination of I-309, SYWC and kallistatin served as the most promising combination to predict TB status for the whole dataset reaching the Triage TPP suggested by the WHO. However, this was reached with only one out of four algorithms applied. Figure 2 clearly shows that adding more biomarkers leads to more information for the algorithms and thus similar behavior in their ROC curve outcomes indicating stability towards the interpretation of the results.

We proceeded to exclude I-309 from the analysis given the difficulty in combining it in an assay with other biomarkers due to its vastly different dynamic range (substantially lower absolute concentration, see Table S4 in the Appendix). In this analysis, SYWC showed up as the best performing single biomarker (AUC 0.86 for logistic regression) closely followed by MIG (AUC 0.83 for logistic regression) and IP-10 (AUC 0.79 for logistic regression). Quantitative information on the host markers’ concentration and fold-changes at global and regional level can be found in Table S4 – S7 in the Appendix.

For biomarker combinations excluding I-309, the two biomarker combination of SYWC and kallistatin was the most promising (AUC 0.88 for logistic regression). Combining more than three biomarkers did not substantially improve accuracy of the prediction towards the TPP (see Appendix Figure S2).

In comparison to the WHO endorsed biomarker CRP as a screening test, the best performing single biomarker, I-309 (AUC 0.87 for logistic regression) alone, outperformed CRP (AUC 0.72 for logistic regression) in our analysis for the whole dataset. At a specificity of 70%, I-309 showed 81% sensitivity for logistic regression, whereas CRP obtained only 75% sensitivity. Considering the better performing support vector machines algorithm, the difference was 89 % sensitivity for I-309 compared to 74 % for CRP. Detailed ROC plots for CRP only are presented in the supplementary material in Figure S1.

### Stratification by country and HIV status

When assessing the data by country, the single biomarkers and all of their combination performed substantially worse in the participants from Vietnam than the other two countries (Figure 3 a) – c)). When excluding Vietnam, the minimal requirements of the TPP were reached for all 12 biomarkers combined with all the machine learning algorithms except Naïve Bayes, as seen in the ROC plots for the remaining dataset (AUC 0.92 with logistic regression excluding Vietnam vs 0.89 with all data combined Figure 3d) and e). Furthermore, the top performing markers identified at the global level (I-309 and SYWC) were among the best performing markers at country level in South Africa and Vietnam. In Peru, SYWC was followed by CRP and I-309. For the other biomarker combinations there are great overlaps of the global level with each country level. See Table 1 and Table S9 in the supplementary material for details.

For HIV positive patients, the combination of SYWC and I-309 reached minimal target accuracy of the TPP using SVM and Random Forest (AUC 0.90 and 0.85, respectively), however the sample size was small (110). No other biomarker subset performed substantially better in HIV positive or negative patients. The stratification by HIV status revealed that with three or more biomarkers performance did not improve in both stratification groups. When excluding I-309, the stratification by HIV status leads to similar results as in the full dataset.

## Discussion

In this multi-center retrospective nested cohort study, several host biomarkers alone and a combination of two to three biomarkers were substantially better than CRP alone, the screening test currently recommended by WHO. A signature of host biomarkers met the minimum WHO criteria for a triage test in adults from two countries (Peru and South Africa), and separately in HIV-positive patients.

The combination I-309, SYWC and kallistatin served as the most promising combination to predict TB disease for the whole dataset. These top-performing biomarkers are part of three different signaling pathways, with a complementary nature. While I-309 stimulates chemotaxis of monocytes and is secreted by activated T lymphocytes (26-28), SYWC is a gamma interferon-inducible Trp-tRNA-synthetase associated with stress response (29) and kallistatin is an endogenous human serine proteinase inhibitor, that is able to inhibit tissue kallikrein kininogenase and amidolytic activities in vitro (30). It is worthwhile to note that kallistatin goes in opposite directions compared to SYWC and I-309; I-309 and SYWC are upregulated in TB patients while kallistatin is down regulated. This might contribute to control for both pre-analytical and analytical sources of variation under the assumption that both proteins undergo the same effects of pre-analytical and analytical variation in the same sample. This also allows for self-normalization and might amplify the signal-to-noise ratio (31). This concept has been applied in other commercial tests (32).

The results were confirmed by different bioinformatics approaches and the most significant and robust biomarkers were identified across different black box algorithms. This analysis aimed at providing a potential set of host markers that could serve as predictors for TB and inform a future research agenda towards the development of a host protein biomarker TB triage test. The study benefitted from prior work that screened a large number of possible markers and helped to select the most promising (8, 18-20) ones.

This lends hope to the feasibility of a simple low-cost, blood-based host protein biomarker assay as currently available multiplex point-of-care assays for antigens and cardiac biomarkers do include three targets, with the latter often being quantitative. In the context of a TB biomarker signature, the assay would need to be quantitative, preferably present a large dynamic range given the concentration of the biomarker candidates and be linked with some processing unit to compute and display the associated TB call, all these challenges could be addressed by recent fluorescent multiplex immunoassays that run on small and portable devices. Whether such an assay is possible at the <2USD target ex-works price at scale remains to be proven (6). Defining a cut-off for a globally applicable test might be difficult, given the variability between regions as seen in our data.

The decreased performance of the signature in Vietnam was notable, and could relate to the patient population enrolled (despite inclusion criteria being the same), other concomitant infections, the host immune system, the circulating *mycobacterium tuberculosis* lineages, or to preanalytical or storage issues of the sample (33, 34). Differences in the population enrolled are suggested by a larger percentage of smear-negative TB. The impact of the host on acute phase reactants was suggested by others, with the Asian ethnicity being associated with lower median baseline pre-treatment CRP (35). This was also confirmed in our data by CRP response being substantially lower in the Vietnam subset. The preanalytical factors and storage issues appear less likely as proteins are expected to be largely stable, sample were recently collected and preanalytical steps were standardized. However, the host and pathogen variability or an interaction of the two could be explanatory. More data are necessary to validate this finding. If this is indeed due to host factors, then such host marker-based tests are necessary to serve a regional market, which is possible but given the low margin on TB diagnostic tests even less attractive for commercialization.

The small sample size of the HIV positive group in the dataset does not allow for general conclusions, in particular further stratification by country was not possible. Furthermore, an evaluation of these biomarkers in children would be useful to assess the added value in this population, where overall diagnostic capabilities are limited and a test on an easily accessible sample (e.g., blood from a finger prick) is urgently needed.

Our study has several limitations that are noteworthy. First of all, we are aiming to define biomarker combinations suitable for a possible triage test, however, the population used (facility-based), was not representative of a population that would be reached with a community-based triage test, as prevalence was very high. It is very likely that in patients presenting earlier in the disease the performance of an algorithm would be worse. Secondly, our population was limited in that it did not include regions with high prevalence of other parasitic diseases and additional host genome variability as would be expected in sites for example in South Asia and equatorial Africa. Thirdly, while we were trying to utilize standardized samples from the FIND biobank, possible pre-analytical or storage issues only affecting one site (e.g. Vietnam) could also potentially explain variability observed in the results. Fourth, excluding patients with unclear microbiological results from our sputum-based reference standard could have led to bias in the performance assessment of our non-sputum-based blood signatures.

In conclusion, a host protein biomarker assay is feasible in adults for certain geographical regions based on one or two biomarkers with a performance that meets minimal WHO TPP criteria (i.e. single marker I-309; or combination of I-309, SYWC and kallistatin to leverage the benefits for assay development outlined above; or I-309 and CRP to leverage existing recommendations). However, more work is needed to demonstrate that such as assay can be translated into a practical point-of-care test, and to better understand how to address regional differences in biomarker levels and responses.

## Data Availability

All data produced in the present study are available upon reasonable request to the authors

## Acknowledgement

We are thankful for the data received from Pham Ngoc Thach Hospital in Ho Chi Minh City, Vietnam (PNTH) as valuable contribution for this evaluation.

