## Appendix for "Diagnostic performance of host protein signatures as a triage test for active pulmonary TB"

*Figure S1: Performance of ApoA1 and CRP individually*

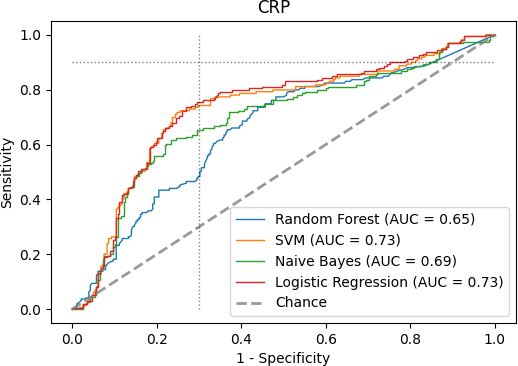

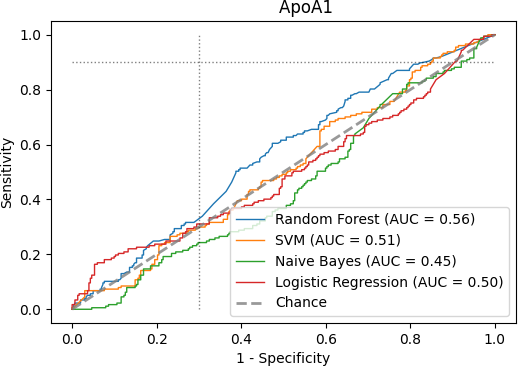

Caption: The dotted line indicates the TPP with 90% sensitivity and 70% specificity. Abbreviations: SVM: Support Vector Machine; AUC: area under the curve

*Figure S2: When excluding I-309, taking more than 3 biomarkers did not substantially improve accuracy of the prediction towards the TPP.*

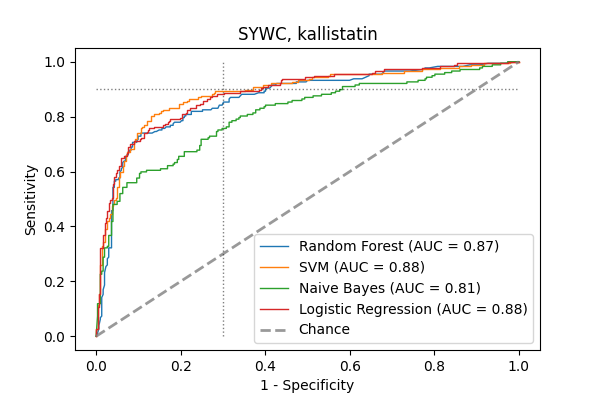

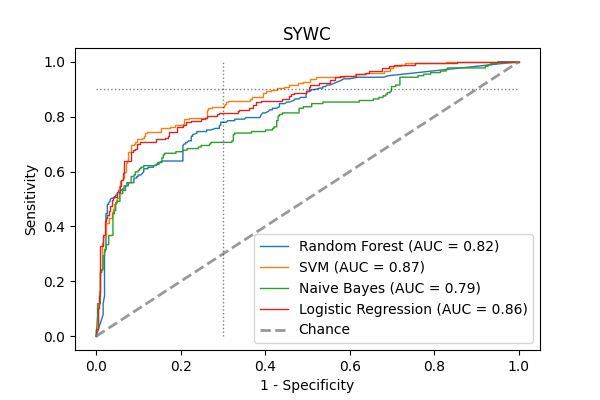

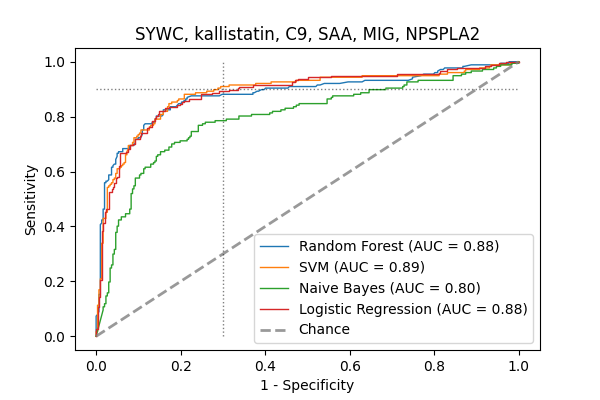

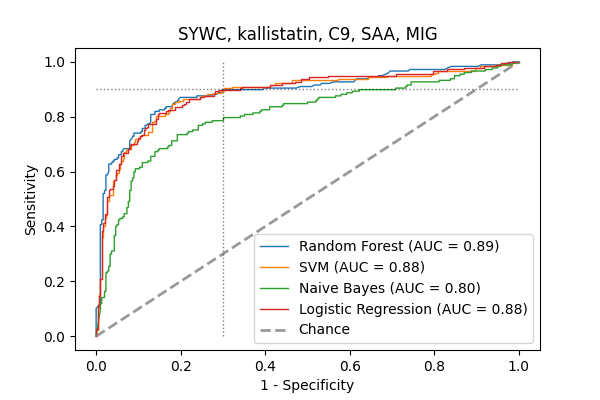

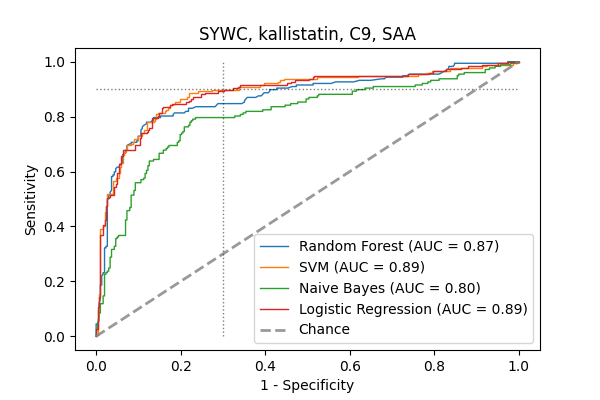

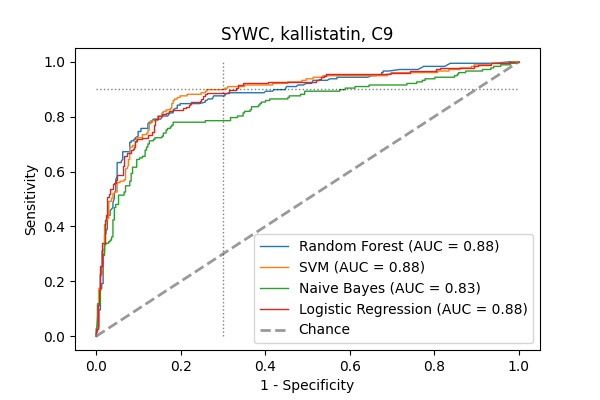
*Figure S3: Most promising biomarker combination for HIV positive patients reaching minimal target accuracy of the TPP with all algorithms except Naïve Bayes. The sample size is 110.*

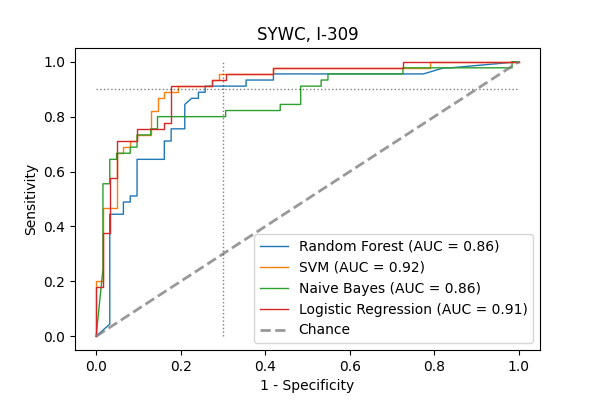

*Table S4:* *Quantitative data on concentration and fold-changes of the single host markers at the global level*

| Host marker | Quantative value  TB positives  Mean, [25th and 75th percentiles] | Quantative value  TB negative  Mean, [25th and 75th percentiles] | Fold change  TB negative/  TB positive  Mean, [25th and 75th percentiles] |
| --- | --- | --- | --- |
| NPS-PLA2 | 244205  [23340, 262547] | 100354  [10250, 38946] | 0.41  [0.44, 0.15] |
| SYWC | 201565  [74824, 245329] | 60320  [32178, 67047] | 0.3  [0.43, 0.27] |
| C9 | 87393926  [58647252, 104369085] | 58914995  [34953155, 69637726] | 0.67  [0.6, 0.67] |
| LBP | 10638841  [4960943, 12109847] | 5779719  [2566144, 6192901] | 0.54  [0.52, 0.51] |
| CRP | 184468404  [44442307, 260598048] | 67569950  [2747640, 44603133] | 0.37  [0.06, 0.17] |
| SAA | 270013188  [22332527, 301088370] | 131027361  [2291012, 29324382] | 0.49  [0.1, 0.1] |
| Kallistatin | 16858488  [10120463, 20992742] | 26393738  [17787049, 30075939] | 1.57  [1.76, 1.43] |
| Ferritin | 640926  [175864, 735009] | 342358  [72881, 416977] | 0.53  [0.41, 0.57] |
| ApoA1 | 2137295290  [626983785, 1542702584] | 10841296498  [977186634, 1921571968] | 5.07  [1.56, 1.25] |
| IP-10 | 1958  [487, 2348] | 571  [173, 599] | 0.29  [0.36, 0.25] |
| I-309 | 126  [36, 100] | 24  [15, 27] | 0.19  [0.41, 0.27] |
| MIG | 6266  [2082, 6809] | 1558  [585, 1704] | 0.25  [0.28, 0.25] |

*Table S5: Quantitative data on concentration and fold-changes of the single host markers in Peru for the top 3 performing host markers*

| Host marker  Ranking | Quantative value  TB positives  Mean, [25th and 75th percentiles] | Quantative value  TB negative  Mean, [25th and 75th percentiles] | Fold change  TB negative/  TB positive  Mean, [25th and 75th percentiles] |
| --- | --- | --- | --- |
| Best: SYWC | 165617,  [75509, 210373] | 32060,  [21220, 31858] | 0.19  [0.28, 0.15] |
| 2^nd^ best: CRP | 215182996,  [69348432, 289583579] | 13990406,  [2853470, 13741375] | 0.07  [0.04, 0.05] |
| 3^rd^ best: I-309 | 86,  [47, 92] | 22,  [13, 25] | 0.26,  [0.29, 0.27] |

*Table S6: Quantitative data on concentration and fold-changes of the single host markers in South Africa for the top 3 performing host markers*

| Host marker  Ranking | Quantative value  TB positives  Mean, [25th and 75th percentiles] | Quantative value  TB negative  Mean, [25th and 75th percentiles] | Fold change  TB negative/  TB positive  Mean, [25th and 75th percentiles] |
| --- | --- | --- | --- |
| Best: I-309 | 177,  [47, 162] | 23,  [14, 24] | 0.13,  [0.3, 0.15] |
| 2^nd^ best: SYWC | 248059,  [137744, 308120] | 77205,  [44388, 89431] | 0.31,  [0.32, 0.29] |
| 3^rd^ best: MIG | 6800,  [3133, 8563] | 1626,  [439, 1934] | 0.24,  [0.14, 0.23] |

*Table S7: Quantitative data on concentration and fold-changes of the single host markers in Vietnam for the top 3 performing host markers*

| Host marker | Quantative value  TB positives  Mean, [25^th^ and 75^th^ percentiles] | Quantative value  TB negative  Mean, [25^th^ and 75^th^ percentiles] | Fold change  TB negative/  TB positive  Mean, [25^th^ and 75^th^ percentiles] |
| --- | --- | --- | --- |
| Best: SYWC | 249225,  [68864, 273974] | 65683,  [41459, 72606] | 0.26,  [0.6, 0.27] |
| 2^nd^ best: I-309 | 135,  [ 34, 75] | 27,  [17, 31] | 0.2  [0.5, 0.42] |
| 3^rd^ best: IP-10 | 2052,  [500, 2020] | 644,  [336, 647] | 0.31  [0.67, 0.32] |

*Table S8: Definition of TB status*

| Description | TB status |
| --- | --- |
| Positive MTB culture | Definite tuberculosis  (smear pos / smear neg) |
| Negative MTB culture, negative smear and response to TB treatment | Clinical tuberculosis |
| Smear negative, Xpert and culture negative on all sputum samples and exhibition symptom resolution in the absence of tuberculosis treatment at the 2–3-month follow-up visit | Non-tuberculosis disease |

*Table S9:* Biomarker combinations ranked according to their value of the negative loglikelihood stratified by country

| No of biomarkers combined | **Best combination**  [marker(s)] Loglikelihood, AUC for logistic regression | **Second best combination**  [marker(s)] Loglikelihood, AUC for logistic regression | **Third best combination**  [marker(s)] Loglikelihood, AUC for logistic regression |
| --- | --- | --- | --- |
| *South Africa* |  |  |  |
| 1 | [I-309] 60, 0.89 | [SYWC] 66, 0.90 | [MIG] 73, 0.89 |
| 2 | [SYWC, I-309]  51, 0.93 | [C9, I-309]  52, 0.90 | [kallistatin, I-309]  55, 0.89 |
| 3 | [SYWC, I-309, C9]  47, 0.93 | [I-309, C9, SAA]  48, 0.91 | [ I-309, SYWC, SAA]  50, 0.93 |
| *Peru* |  |  |  |
| 1 | [SYWC] 26, 0.88 | [CRP] 29, 0.88 | [I-309] 29, 0.90 |
| 2 | [SYWC, C9]  40.1, 0.92 | [ApoA1, CRP]  40.5, 0.88 | [I-309, SYWC]  40.6, 0.92 |
| 3 | [SYWC, I-309, ApoA1]  19, 0.93 | [SYWC, CRP, ApoA1]  19, 0.91 | [SYWC, C9, ApoA1]  20, 0.92 |
| *Vietnam* |  |  |  |
| 1 | [SYWC] 74, 0.84 | [I-309] 82, 0.82 | [IP-10] 89, 0.76 |
| 2 | [SYWC, Ferritin]  73, 0.84 | [SYWC, I-309]  74, 0.84 | [SYWC, kallistatin]  74, 0.83 |
| 3 | [SYWC, I-309, Ferritin]  72, 0.84 | [SYWC, Ferritin, kallistatin]  73, 0.83 | [SYWC, kallistatin, SAA]  73, 0.83 |

*S10*: Additional information regarding MSD U-PLEX assay testing

For each host biomarker, antibodies were first screened to identify suitable antibody pairs when not previously identified by MSD (kallistatin, NPS-PLA2, SYWC, ferritin) and single-plex testing was conducted for calibrator titration and native protein recognition in human serum. Then, feasibility of multiplexing and further qualification of the panels were performed by collecting data on cross-reactivity between the members of the panel, calibrator curves, dilution linearity in diluent, spike recovery in diluent, reproducibility and finally stability of the host biomarkers in human serum.

Each plate run during sample testing included a set of eight calibration samples (created by a 1:4 serial dilution of the assay calibration standard), the three controls and up to 37 serum samples, all run in duplicate. The assays were calibrated by fitting the relationship of the calibration sample signals to their assigned concentrations with a four-parameter logistic (4-PL) model using 1/Y^2^ weighting. Quantitation of the samples and controls was carried out by back-fitting the assay signals to the 4-PL model, and then correcting for the sample dilution.

The custom assay panels were created using antibodies pairs that are available commercially from MSD as R-PLEX^®^ or U-PLEX antibody sets, with the exception of the NPS-PLA2 and SYWC assays, which used prototype antibody pairs selected specifically for this study. The custom panels were qualified through verification studies characterizing cross-reactivity between the assay targets in each panel, dilution linearity in diluent, spike recovery in diluent, reproducibility and stability of the host biomarkers in human serum.
